# Association between Quality of Antenatal Care and Postpartum Depression among Women in Al Anbar Governorate, Iraq: A Cross-Sectional Study

**DOI:** 10.1101/2025.08.12.25333535

**Authors:** Ruya Abdulhadi M. Saeed, Nibras Alaa Hussain

**Affiliations:** University of Anbar, College of Medicine/Dept. of Community and Family medicine, Ramadi, Iraq; University of Nahrain, College of Medicine /Dept. of Community and Family Medicine, Baghdad, Iraq

**Author notes:** Corresponding author: Ruya Abdulhadi M.Saeed.

**Keywords:** Postpartum depression, Antenatal care quality, Maternal mental health, Conflict-affected settings, Primary Care Center

## Abstract

**Background:** Postpartum depression (PPD) is a leading cause of maternal morbidity, particularly in low-resource and conflict-affected regions. While high-quality antenatal care (ANC) may offer support against maternal mental distress, the association between PPD and ANC remains underexplored in conflict-affected settings.

**Objective:** To investigate the association between perceived quality of ANC and the prevalence of PPD among postpartum women in Al Anbar governorate, Iraq.

**Methods:** In a cross-sectional study conducted between February 2024 and February 2025, 618 women ≤8 weeks postpartum were recruited from 23 primary health care centers (PHCCs) across Al Anbar. ANC quality was assessed using the 46-item Quality of Prenatal Care Questionnaire (QPCQ), and PPD symptoms were measured using the Edinburgh Postnatal Depression Scale (EPDS, cutoff ≥13). Associations between ANC quality and PPD were analyzed using chi-square tests and Pearson correlations.

**Results:** Overall, 42.4% of women screened positive for probable PPD. While 58.1% of participants rated their ANC as “good” (QPCQ ≥165), perceived ANC quality did not significantly correlate with total EPDS scores (r = 0.067, p = 0.094). Subscale analysis revealed that women with PPD paradoxically reported higher scores in information sharing, anticipatory guidance, and service availability. Higher ANC visit frequency and maternal education were significantly associated with higher QPCQ scores, yet these did not correspond to lower PPD prevalence. Geographic differences in ANC quality were observed but did not align with PPD incidence.

**Conclusions:** In conflict-affected Al Anbar, higher perceived ANC quality did not translate into reduced PPD risk. These findings highlight the need for integrated psychosocial support within ANC programs and stress the limitations of standard prenatal care in addressing maternal mental health in conflict-affected contexts.

## Introduction

Postpartum depression (PPD) is a serious global public health concern, affecting an estimated 15–20% of new mothers worldwide. Unlike the transient “baby blues,” PPD encompasses persistent feelings of sadness, hopelessness, irritability, and, in severe cases, thoughts of self-harm or harm to the infant, severely impairing maternal functioning and mother–child bonding (Cox et al., 1987; Gjerdingen & Yawn, 2007). Although the prevalence of PPD is variably reported, it is estimated at around 17.2% globally and up to 19.8% in low- and middle-income countries (Fisher et al., 2012), though the true burden is likely underdetected due to limited screening in many regions (Alwan et al., 2018; Roberts et al., 2009). Early detection and timely psychosocial or pharmacological intervention are key to mitigating these adverse outcomes, yet stigma and limited mental health resources often impair treatment—especially in resource-constrained settings (Tol et al., 2011; McFarlane et al., 2011).

Comprehensive antenatal care (ANC) extends beyond routine obstetric monitoring to encompass health education, psychosocial support, and the early identification of mental-health risks. High-quality prenatal services, characterized by clear information sharing, respectful communication, anticipatory guidance, and accessible resources, have been shown in stable settings to mitigate PPD risk by promoting maternal empowerment and trust in the health system (Heaman et al., 2015; Sword et al., 2012). Yet in many under-resourced contexts, mental-health screening and counseling remain scarce or absent, leaving expectant women more vulnerable to unrecognized distress (Ismail et al., 2016; Thabet & Thabet, 2019).

Al Anbar governorate in western Iraq presents a particularly challenging environment for maternal care. Years of conflict and displacement have stressed primary health care centers (PHCCs), resulting in shortages of trained staff, intermittent supply chains for medicines and equipment, and uneven distribution of services between urban hubs like Ramadi and outlying districts (Ministry of Health Iraq, 2023). Insecurity and infrastructure damage hamper outreach to peripheral communities, where distance, transport barriers, and shifting population movements further limit access to even basic antenatal visits (WHO Iraq, 2022).

Against this healthcare system fragility and elevated psychosocial stressors, understanding how perceived ANC quality relates to PPD is vital. Investigating this association in Al Anbar not only fills a gap in conflict-affected settings but also informs context-specific interventions, such as integrating routine mental-health screening into prenatal visits (WHO, 2013), strengthening staff training in perinatal psychosocial support, and tailoring resource allocation to underserved districts (Betancourt et al., 2010). This study aims to quantify the link between women’s perceptions of antenatal care quality and the prevalence of PPD across Al Anbar’s diverse PHCC network, with the ultimate goal of guiding policy and programmatic efforts to safeguard maternal mental health in high-adversity regions.

## Materials and Methods

### Study Design and Setting

A cross-sectional survey was conducted between February 2024 and February 2025 across 23 PHCCs in Al Anbar governorate, Iraq. Centers were selected via simple random sampling, covering both urban and peripheral districts.

### Ethical Approval

Ethical clearance was obtained from the Institutional Review Board of Al-Nahrain College of Medicine. Permissions were also secured from the Directorate of Health (DOH) in Anbar. Written informed consent was obtained from all participants, ensuring confidentiality and voluntary participation, in accordance to the declaration of Helsinki.

### Participants

Mothers up to 8 weeks postpartum, attending PHCCs for postnatal care or infant vaccination, were consecutively invited. Exclusion criteria included pre-existing major psychiatric disorders, refusal to participate, and incomplete interviews.

### Measures

Quality of Prenatal Care Questionnaire (QPCQ): A validated 46-item tool with six subscales (information sharing, time and availability, decision guidance, approachability, respect and support, and availability of services) scored 44–220. Scores ≥165 denote good antenatal care quality (Sword et al., 2012).

Edinburgh Postnatal Depression Scale (EPDS): A 10-item questionnaire assessing depressive symptoms over the past week, scored 0–30; a cutoff ≥13 indicates probable PPD (Cox et al., 1987).

Sociodemographic data: Age, parity, education, income, and social support were collected via structured interview.

### Statistical Analysis

Data analysis used SPSS v26. Descriptive statistics summarized participant characteristics. Associations between antenatal care quality (good vs. poor) and PPD were assessed with Pearson’s chi-square test. Odds ratios (ORs) and 95% confidence intervals (CIs) were calculated. Significance threshold: p<0.05.

## Results

A total of 618 women were enrolled in the study. The mean age of the mothers was 28.4±5.7 years, with the youngest participant being 16 and the oldest 46. Nearly 15% of mothers (n=91) were adolescents under 20 years old, slightly over half (n=323; 52.3%) fell into the 20–29-year category, 28.2% (n=174) were aged 30–39 years, and only 4.9% (n=30) were aged 40 or more. Their partners tended to be older: 43.7% of husbands (n=270) were under 30, 55.7% (n=344) were between 30 and 49, and a very small minority (n=4; 0.6%) were 50 years or above.

Regarding postpartum follow-up, most mothers (63.4%; n=392) were assessed within the first four weeks after delivery, while the remaining 36.6% (n=226) were evaluated between four and 8 weeks. In terms of residence, nearly two-thirds of participants (63.9%; n=395) lived in urban centers, and 36.1% (n=223) resided in more peripheral or rural regions. Household composition varied: just over half of the women (54.2%; n=335) lived with their husband’s extended family, whereas 45.8% (n=283) remained in their own nuclear family home.

### Socioeconomic and Educational Profile

The vast majority of mothers (72.7%; n=449) were homemakers, with the remaining 27.3% (n=169) engaged in paid employment. In contrast, most husbands (77.7%; n=480) were employed, and 22.3% (n=138) reported no current occupation. Economic status was reported as sufficient in 71.4% of families (n=441), while 28.6% (n=177) had an insufficient income.

Education lavels also showed a varied dataset. Among mothers, 9.5% (n=59) were illiterate, 35.6% (n=220) had completed only primary school, 26.9% (n=166) had a secondary education, 18.6% (n=115) held an undergraduate degree, and 9.4% (n=58) had pursued postgraduate studies. Husbands’ educational levels were similarly distributed: 9.7% (n=60) were illiterate, 26.4% (n=163) had primary schooling, 29.9% (n=185) secondary, 23.1% (n=143) academic, and 10.8% (n=67) postgraduate.

### Antenatal Care Quality is increased by higher visit frequency and maternal education

Women who attended regular ANC visits reported significantly higher perceived care quality, with a mean QPCQ score of 144.59 (SD = 25.31) compared to 133.82 (SD = 30.69) among those with irregular visits (t(616) = 4.78, p < 0.001). Maternal education was significantly associated with antenatal care quality (χ^2^ = 13.50, p = 0.009). The highest proportion of “good” QPCQ ratings (≥165) occurred in women with secondary education (57.8%), followed by those with primary schooling (42.7%), undergraduate degrees (43.5%), and illiterate mothers (40.7%).

### Prevalence of Postpartum Depression and Antenatal Care Quality

Overall, 42.4% of mothers (n=262) scored ≥13 on the Edinburgh Postnatal Depression Scale, indicating probable postpartum depression, while 57.6% (n=356) scored below this threshold. Antenatal care quality, measured by QPCQ score, revealed that globally 58.1% of women (n=359) rated their care as “good” (QPCQ total ≥165), whereas 41.9% (n=259) considered it “poor.” A bivariate Pearson correlation revealed a very weak, non-significant positive relationship between total QPCQ and EPDS scores (r = 0.067, p = 0.094), suggesting that overall prenatal care quality alone does not linearly predict depressive symptoms. Surprisingly, further analysis of the six QPCQ subscales revealed that information sharing in mothers with PPD recieved higher scores compared to those without PPD (PPD = 28.4, SD = 6.4; no PPD = 27.2, SD = 6.4; t = –2.11, p = 0.036). In addition, anticipatory guidance (no PPD = 33.3, SD = 8.5; PPD = 35.0, SD = 8.1, t = –2.46, p = 0.014) and availability (PPD = 16.2, SD = 4.1; no PPD = 15.4, SD = 4.0); t = –2.21, p = 0.027) also showed higher values, therefore a better quality of care, between women with PPD, as compared to women without such diagnosis. The other three aspects, sufficient time, approachability and support&respect did show any differences between the two groups.

### Geographic Variations of prenatal care and PPD incidence

We also examined district-level differences in antenatal care, which were not related to the urban or rural background of the PHCC: in the largest urban centers, in fact, we observed a generally high antenatal score, with 67% prevalence of good quality in Falluja and 62% in Ramadi, while other cities like Heet showed a low 40.9% of women served with good prenatal care. The rural areas varied widely from the 54.5% of Al Rutba to the surprising 80% of percieved good quality care in Ana. Finally, the significiant differences across district did not correlate with the levels of PPD, where we observed high levels of PPD, 52%, in Ramadi first, where the antenatal care was within the highest, and observed also low levels of PPD in disctricts with low quality of care, like in Ramadi second, where the low low incidence of PPD (37%) is in contrast with the very low 37% of perceived good quality care (Table 1).

**Table 1.** Geographical differences in precieved antenatal the Al Anbar district, coupled with the prevalence of PPD in the same areas.

| Code | Total | Bad quality (< 155)<br>(n=333) | Good Quality (≥155)<br>(n=285) | PPD prevalence (%) |
| --- | --- | --- | --- | --- |
| Al- Ramadi First | 79 | 30 (38.0%) | 49 (62.0%) | 51.9 |
| Fallujah | 97 | 32 (33.0%) | 65 (67.0%) | 43.3 |
| Al- Ramadi Second | 73 | 46 (63.0%) | 27 (37.0%) | 37 |
| Heet | 44 | 26 (59.1%) | 18 (40.9%) | 63.6 |
| Al- Khalidia | 56 | 19 (33.9%) | 37 (66.1%) | 28.6 |
| Al- Rutba | 22 | 10 (45.5%) | 12 (54.5%) | 31.8 |
| Al- Karma | 38 | 13 (34.2%) | 25 (65.8%) | 42.1 |
| Al- Qa'im | 47 | 19 (40.4%) | 28 (59.6%) | 42.6 |
| Haditha | 68 | 26 (38.2%) | 42 (61.8%) | 38.2 |
| Al-Faris | 40 | 22 (55.0%) | 18 (45.0%) | 42.5 |
| Ana | 32 | 6 (18.8%) | 26 (81.2%) | 40.6 |
| Al- Baghdadi | 22 | 10 (45.5%) | 12 (54.5%) | 40.9 |

## Discussion

Our analysis revealed that regular antenatal-care (ANC) visits were significantly associated with better perceived quality of prenatal services. Women with secondary education reported the highest prevalence of “good” antenatal care (57.8%), a pattern consistent with European and Japanese studies linking maternal literacy to more effective guidance and patient support (Tocchioni et al., 2018, Iida et al., 2012).

Moreover, mothers attending regular ANC visits scored higher on the QPCQ than those with irregular visits, underscoring continuity of care as a key quality driver.

We found that the overall correlation between total QPCQ score and EPDS was weak and non-significant, and we detected that higher scores for information sharing, anticipatory guidance, and availability of services were significantly associated with higher incidence of PPD symptoms, whereas sufficient time, approachability, and support & respect were not. This analysis highlights two key points: first, higher ANC visit frequency and greater maternal education are associated with better perceived prenatal care quality; second, better perceived care quality does not translate into reduced PPD symptoms in this conflict-affected setting. Moreover, certain QPCQ subscales are rated more highly by women with PPD, suggesting complex dynamics in the tested areas.

Our finding that ≥4 ANC visits and higher education predict superior QPCQ ratings aligns with literature showing that frequent contact promotes interpresonal connection and that educated women may be more empowered and able to actively seek for comprehensive care (Heaman et al., 2015; Sword et al., 2012). In stable settings, these factors often correspond to improved maternal outcomes, yet here they did not protect against PPD. The absence of a protective effect of perceived antenatal care quality on PPD is in contrast with numerous studies in non-conflict contexts (Heaman et al., 2015; Sword et al., 2012). In war-afflicted regions, however, pervasive trauma, insecurity, and resource disruption may overwhelm the psychosocial benefits of even high-quality prenatal services (Roberts et al., 2009; Thabet & Thabet, 2019). Thus, while women value information and availability, these interventions alone may be insufficient to buffer against conflict-induced stress.

Our finding,in fact, highlight a 42.4% prevalence of probable PPD (EPDS ≥13) in Al Anbar, higher than most global estimates (10–20%) but broadly consistent with rates observed in other conflict-affected or low-resource settings. For example, a survey of Syrian refugee women in Lebanon reported a 45.2% PPD prevalence, attributing elevated risk to trauma exposure and disrupted health services (Alwan et al., 2018). In contrast, community-based studies in stable middle-income countries tend to report lower rates, like 24.3% in urban Malaysia (Ismail et al., 2016) and 28.7% in rural Ethiopia (Gebremedhn & Gebremariam, 2017), highlighting the amplifying effect of insecurity and displacement on maternal mental health.

Geographic disparities in perceived antenatal care quality across Al Anbar’s PHCC network further emphasize the importance of local health system dynamics. Despite expectations that urban centers would consistently offer superior care, this was not uniformly observed. While Fallujah and Al-Ramadi First reported high proportions of good care (67% and 62%, respectively), other large cities such as Heet showed much lower quality perceptions (40.9%). Conversely, some peripheral or rural districts like Ana reported unexpectedly high quality (81.2%), possibly reflecting either strong individual PHCC performance or lower patient expectations. These findings align with prior research in conflict-affected and resource-limited regions, which suggest that variability in facility-level leadership, staffing, and NGO engagement may outweigh urban–rural distinctions in shaping maternal care quality (Roberts et al., 2009; Betancourt et al., 2010).

Three subscales we analyzed, Information Sharing, Anticipatory Guidance, and Availability, were rated higher by women with PPD, apparently contradicting the usefullness of antenatal care. However, several coherent explanations can be hypothesized: women experiencing or anticipating distress may actively seek more information, resulting in higher self-reported scores (McFarlane et al., 2011). Depressed mothers could overreport service attempts at support, perceiving routine explanations as more relevant (Gjerdingen & Yawn, 2007). Finally, health workers may intensify information and guidance when mothers show emotional distress, generating higher subscale scores in the PPD group.

Similar patterns have indeed been observed in conflict settings: Syrian refugee women with high distress rates reported better perceived counseling efforts, attributed to targeted outreach by NGOs (Alwan et al., 2018). These differences underscore that quantitative care ratings can reflect both provider effort and patient need.

For these reasons, in war-affected Al Anbar, improving perceived ANC quality alone may not suffice to prevent PPD. Integrated models combining prenatal mental health screening, trauma-informed counseling, and community-based psychosocial support are likely needed (Tol et al., 2011; Betancourt et al., 2010).

One of the biggest strenght of this study is that it collected a regionally representative sample across 23 PHCCs and it applied internationaly validated instruments (QPCQ, EPDS). Limitations are related to the experimental design, a cross-sectional study, which does not allow to draw causality from the assocaition detected, and the reliance on self-reported measures potentially susceptible to biases. Selection bias may also arise from convenience sampling at clinic sites. Future longitudinal studies should evaluate whether targeted ANC enhancements (e.g., combined mental-health and education interventions) translate into sustained reductions in PPD incidence in conflict-affected populations.

## Data Availability

All data produced in the present study are available upon reasonable request to the authors, in compliance with the Iraqi laws about confidentialty

## Abbreviations

ANC: antenatal care
EPDS: Edinburgh Postnatal Depression Scale
PHCC: Primary health care center
PPD: post partum depression
QPCQ: Quality of Prenatal Care Questionnaire

## Ethics approval and consent to participate

Ethical approval was obtained from The Institutional review board of Al-Nahrain College of Medicine (IRB), and the permission to collect data from the health sectors of districts was obtained from the Department of Health of Anbar. All patients signed an informed consent prior to inclusion

## Consent for publication

Not applicable

## Availability of data and materials

The datasets generated and/or analysed during the current study are not publicly available due to compliance with patient confidentiality regulations under Iraqi health privacy laws,but can be obtained from the corresponding author upon reasonable request.

## Competing Interests

The authors declare that they have no competing interests

## Funding

This research received no external funding

## Authors’ contributions

Conceptualization, R.A.S. and N.A.H:, methodology, R.A.S. and N.A.H.; formal analysis, R.A.S..; funding and supervision, N.A.H.; data curation and extraction, R.A.S.; original draft preparation, R.A.S.; review and editing, R.A.S. and N.A.H.; project administration, N.A.H.. All authors have read and agreed to the published version of the manuscript.

## Acknowledgements

Not applicable

## Notes

### Competing Interest Statement

The authors have declared no competing interest.

### Funding Statement

This study did not receive any funding

### Author Declarations

Institutional Review Board of Al-Nahrain College of Medicine gave ethical approval

### Summary of Updates

We corrected two references that were incorrect and added two new ones that were left out of the original submission by mistake

